# Weakly supervised learning for multi-organ adenocarcinoma classification in whole slide images

**DOI:** 10.1101/2022.03.28.22273054

**Authors:** Masayuki Tsuneki, Fahdi Kanavati

**Affiliations:** Medmain Research, Medmain Inc., 2-4-5-104, Akasaka, Chuo-ku, Fukuoka, 810-0042, Fukuoka, Japan

**Keywords:** Deep learning, Weakly supervised learning, Adenocarcinoma, Whole slide image, multi-organ

## Abstract

The primary screening by automated computational pathology algorithms of the presence or absence of adenocarcinoma in biopsy specimens (e.g., endoscopic biopsy, transbronchial lung biopsy, and needle biopsy) of possible primary organs (e.g., stomach, colon, lung, and breast) and radical lymph node dissection specimen is very useful and should be a powerful tool to assist surgical pathologists in routine histopathological diagnostic workflow. In this paper, we trained multi-organ deep learning models to classify adenocarcinoma in biopsy and radical lymph node dissection specimens whole slide images (WSIs). We evaluated the models on seven independent test sets (stomach, colon, lung, breast, lymph nodes) to demonstrate the feasibility in multiorgan and lymph nodes specimens from different medical institutions and international public datasets, achieving receiver operating characteristic areas under the curves (ROC-AUCs) in the range of 0.91-0.99.

## 1 Introduction

Adenocarcinoma is a type of carcinoma that has the propensity to differentiate into glandular, ductal, and acinar cells in several organs (e.g., stomach, colon, lung, and breast). According to the Global Cancer Statistics 2020 Sung et al (2021), number of new deaths and % of all sites for stomach, colon, lung, and breast cancers were as follows: 768,793 cases (7.7%) in stomach, 576,858 cases (5.8%) in colon, 1,796,144 cases (18.0%) in lung, and 684,996 cases (6.9%) in breast. Adenocarcinoma is the major cancer arise in these organs, so that adenocarcinoma classification in the primary organs especially on biopsy specimens is one of the most important histopathological inspection in clinical workflow to determine the strategies of cancer treatment. Moreover, lymph nodes are the most common site of metastatic adenocarcinoma, and can be constituted the first clinical manifestation of the cancer. The important clinical practice of the surgical pathologist is to identify the presence or absence of a malignant process in the lymph node. If cancer cells are identified within the efferent lymph vessels and extra-nodal tissues, it is necessary to note in the pathological report because of the possible prognostic significance. Histopathological evaluation of lymph node metastasis is very important for staging of tumors, documentation of tumor recurrence, and prediction of the most probable primary site for a metastatic cancer of uncertain primary site. However, in the routine practical diagnosis, frequently there are numerous number of lymph nodes to be inspected in a single glass slide and there are number of radical lymph node dissection specimen glass slides in the same patient, which should be a workload burden for surgical pathologists.

The incorporation of deep learning models in routine histopathological diagnostic workflow is on the horizon and is a promising technology, allowing the potential of reducing the burden of time-consuming diagnosis and increasing the detection rate of anomalies including cancers. Deep learning has been widely applied in histopathological cancer classification on whole-slide images (WSIs), cellular detection and segmentation, and the stratification of patient outcomes Yu et al (2016); Hou et al (2016); Madabhushi and Lee (2016); Litjens et al (2016); Kraus et al (2016); Korbar et al (2017); Luo et al (2017); Coudray et al (2018); Wei et al (2019); Gertych et al (2019); Bejnordi et al (2017); Saltz et al (2018); Campanella et al (2019); Iizuka et al (2020). Previous works have looked into applying deep learning models for adenocarcinoma classification separately for different organ, such as stomach Iizuka et al (2020); Kanavati and Tsuneki (2021b); Kanavati et al (2021a), colon Iizuka et al (2020); Tsuneki and Kanavati (2021), lung Kanavati and Tsuneki (2021b); Kanavati et al (2021b), and breast Kanavati and Tsuneki (2021a); Kanavati et al (2022) histopathological specimen WSIs. Although these existing models exhibited very high ROC-AUCs for each organ, they cannot classify adenocarcinoma across organs accurately.

In this study, we trained deep learning models using weakly-supervised learning to predict adenocarcinoma in WSIs of stomach, colon, lung, and breast biopsy specimens for primary tumors as well as radical lymph node dissection specimens for metastatic carcinoma using training datasets for stomach, colon, lung, and breast biopsy specimen WSIs without annotations. We evaluated the models on each primary organ biopsy specimen (stomach, colon, lung, and breast) and public datasets (lung and breast) as well as radical lymph node dissection specimens to evaluate presence or absence of metastatic adenocarcinoma, achieving and ROC-AUC from 0.91 to 0.99. Our results suggest that deep learning algorithms might be useful for histopathological diagnostic aids for adenocarcinoma classification in primary organs and lymph node metastatic cancer screening.

## 2 Materials and methods

### 2.1 Clinical cases and pathological records

In the present retrospective study, a total of 8,896 H&E (hematoxylin & eosin) stained histopathological specimen slides of human adenocarcinoma and non-adenocarcinoma (adenoma and non-neoplastic) lesions were collected from the surgical pathology files of five hospitals: International University of Health and Welfare (IUHW), Mita Hospital (Tokyo, Japan) and Kamachi Group Hospitals (total four hospitals: Wajiro, Shinkuki, Shinkomonji, and Shinmizumaki Hospital) (Fukuoka, Japan) after histopathological review by surgical pathologists. The histopathological specimens were selected randomly to reflect a real clinical settings as much as possible. Prior to the experimental procedures, each WSI diagnosis was observed by at least two pathologists with the final checking and verification performed by senior pathologists. All WSIs were scanned at a magnification of x20 using the same Leica Aperio AT2 Digital Whole Slide Scanner (Leica Biosystems, Tokyo, Japan) and were saved as SVS file format with JPEG2000 compression.

### 2.2 Dataset

Hospitals which provided histopathological specimen slides were anonymised (e.g., Hospital-A, B, C, D, and E). Table 1 breaks down the distribution of training sets from four domestic hospitals (Hospital-A, B, C, and D). Table 2 shows the distribution of 1K (1,000 WSIs), 2K (2,000 WSIs), and 4K (4,000 WSIs) training sets. Validation sets were selected randomly from the training sets and the numbers of validation sets were given in parentheses (Table 2). The distribution of test sets from five domestic hospitals (Hospital-A, B, C, D, and E) was summarized in Table 3. In both training and test sets, stomach, colon, lung, and breast WSIs solely consisted of biopsy (stomach and colon: endoscopic biopsy, lung: transbronchial lung biopsy (TBLB), breast: needle biopsy) specimens and lymph node WSIs consisted of radical dissection specimens (Table 1, 2, 3). The distribution of lymph nodes using test sets were summatized in Table 4. All training sets WSIs were not manually annotated and the training algorithm only used the WSI labels which were extracted from the histopathological diagnostic reports after reviewing surgical pathologists; meaning that the only information available for the training was whether the WSI contained adenocarcinoma or non-adenocarcinoma but no information available about the location of the cancerous lesions. In addition to the test sets from clinical institutions, we have used the external lung and breast TCGA datasets as test sets (Table 5) which are publicly available through the Genomic Data Commons (GDC) Data Portal (https://portal.gdc.cancer.gov/). We have confirmed that surgical pathologists were able to diagnose these cases from visual inspection of the H&E stained slide WSIs alone.

**Table 1:**
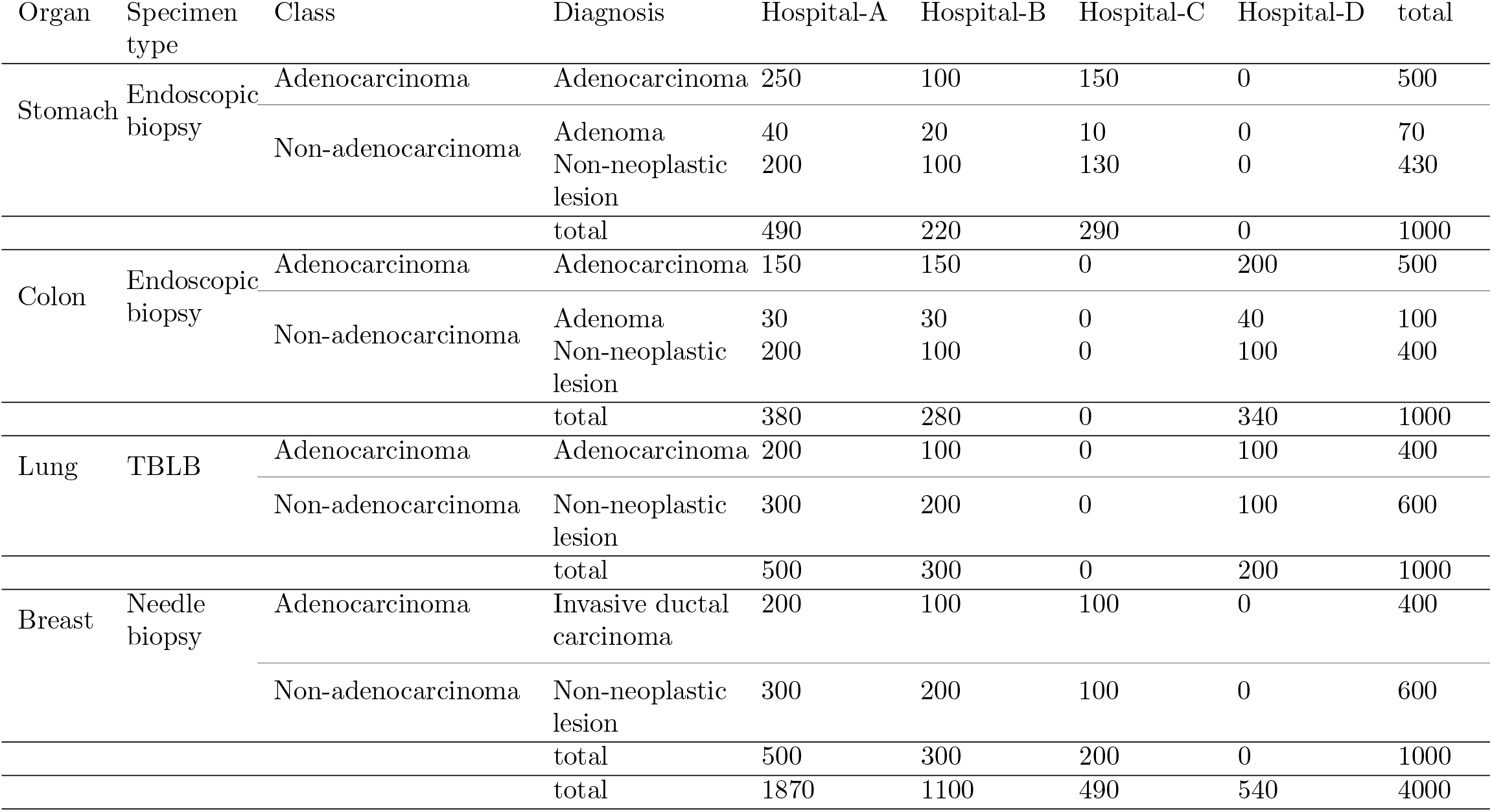
Distribution of cases in the training sets obtained from different hospitals (A-D)

**Table 2:**
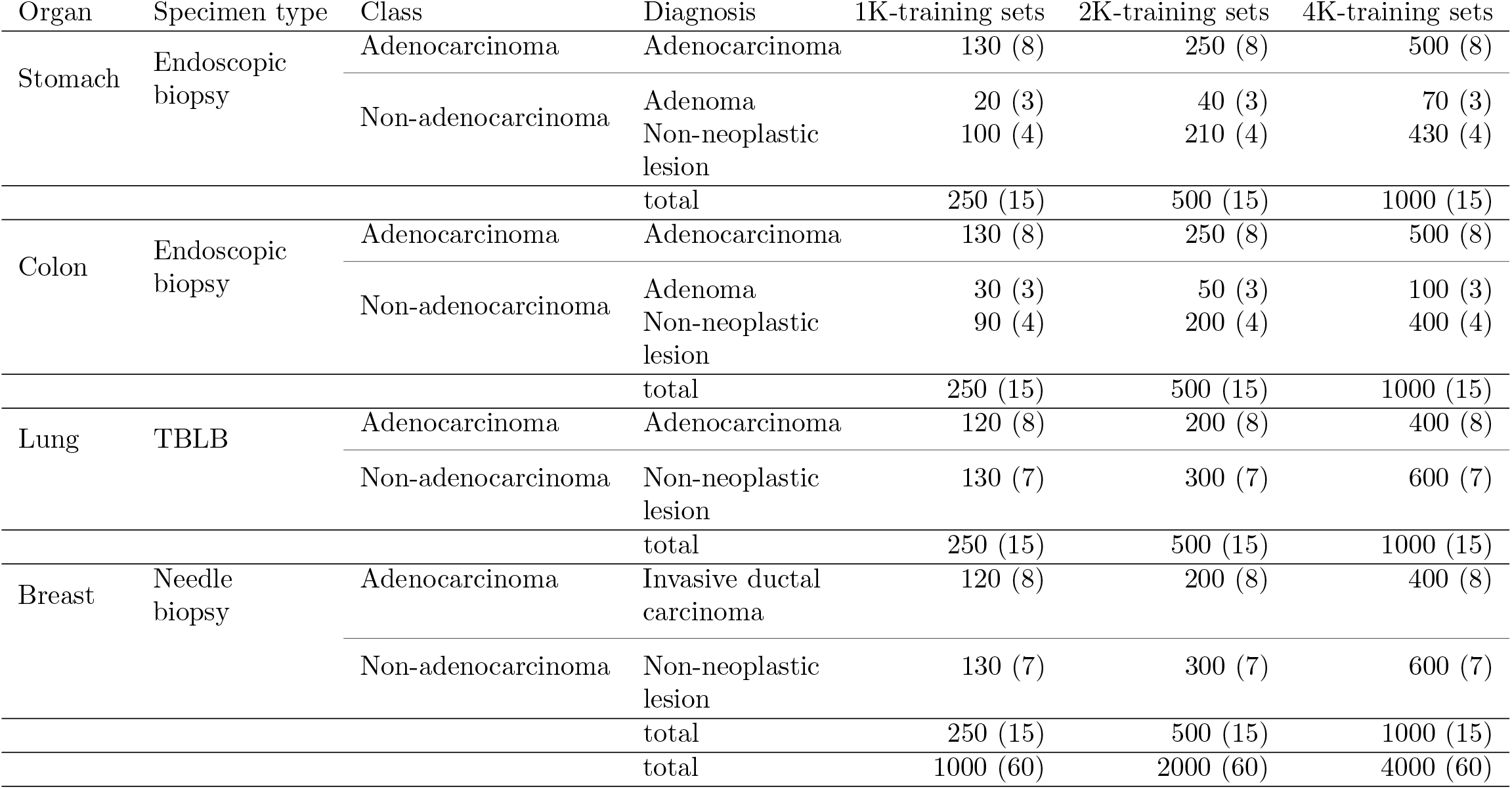
Distribution of cases in the training sets and validation sets. The numbers of validation cases are given in parentheses.

**Table 3:**
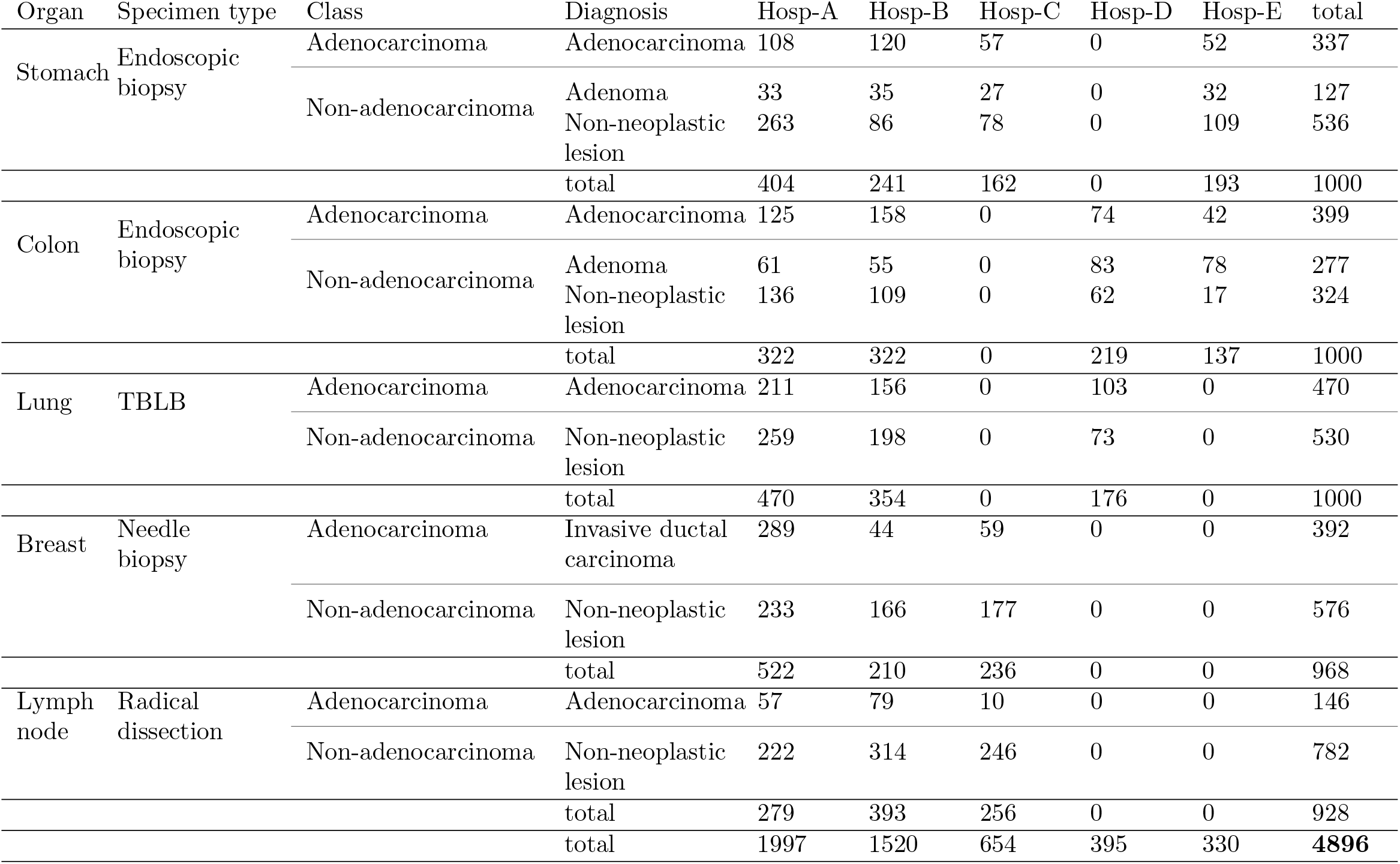
Distribution of cases in the test sets obtained from hospitals (A-E)

**Table 4:** Distribution of whole slide images (WSIs) in the lymph nodes test sets.

| Resected organ | Clinical diagnosis | Histopathological diagnosis | WSI |
| --- | --- | --- | --- |
| Stomach | Advanced gastric cancer | Adenocarcinoma | 18 |
|  |  | Non-neoplastic lesion | 97 |
| Colon | Advanced colon cancer | Adenocarcinoma | 21 |
|  |  | Non-neoplastic lesion | 166 |
| Lung | Lung cancer | Adenocarcinoma | 38 |
|  |  | Non-neoplastic lesion | 181 |
| Lung | Metastatic colon cancer | Adenocarcinoma | 27 |
|  |  | Non-neoplastic lesion | 172 |
| Breast | Breast cancer | Invasive ductal carcinoma | 42 |
|  |  | Non-neoplastic lesion | 166 |

**Table 5:**
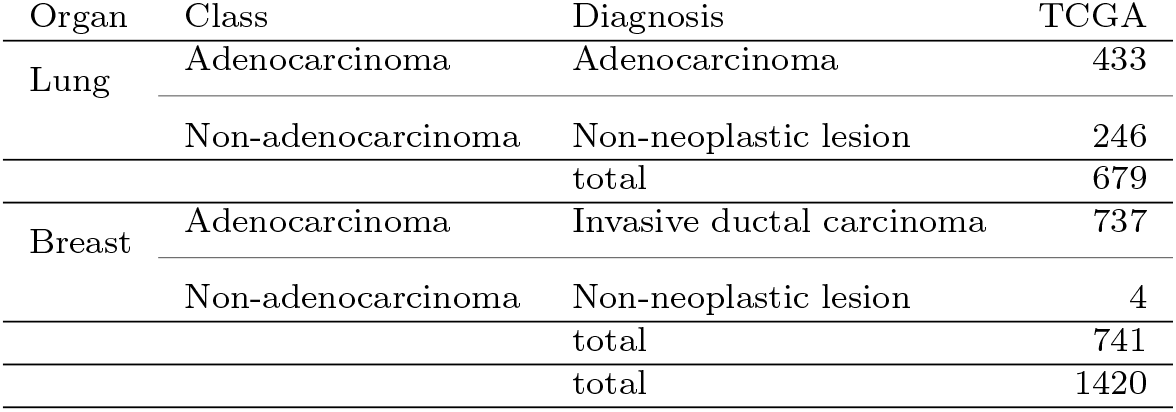
Distribution of cases in the test set obtained from the TCGA public dataset

### 2.3 Deep learning models

In this study, we used the EfficientNetB1 Tan and Le (2019) as the architecture of our models. We use the partial fine-tuning approach Kanavati and Tsuneki (2021c) to train them. This method consists of using only fine-tuning the affine parameters of the batch normalization layers and the final classification layer while leaving the remaining weights of an existing pre-trained model frozen. Figure 1 shows an overview of the training method.

**Fig. 1:**
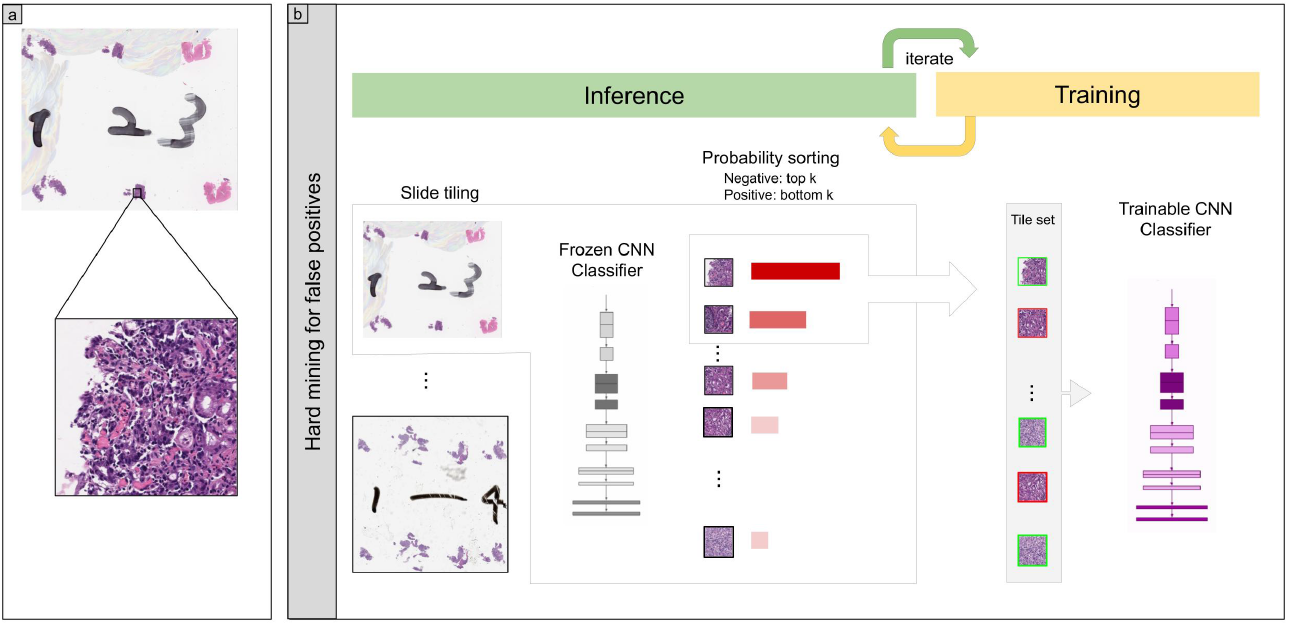
Overview of training method. (a) shows a zoomed-in example of a tile from a WSI. (b) During training, we alternated between an inference step and a training step. During the inference step, the model weights were frozen and the model was used to select tiles with the highest probability after applying it on the entire tissue regions of each WSI. The top k tiles with the highest probabilities were then selected from each WSI and placed into a queue. During training, the selected tiles from multiple WSIs formed a training batch and were used to train the model.

As we only had WSI labels, we used a weakly supervised method to train the models. The training method is similar to the one described in Kanavati et al (2020).

WSIs typically have large areas of white background that is not required for training the model and can easily be eliminated with preprocessing via thresholding using Otsu’s method Otsu (1979). This creates a mask of the tissue regions from which it would then be possible to sample tiles in real-time using the OpenSlide library Goode et al (2013) by providing coordinates from the tissue regions.

### 2.4 Software and statistical analysis

During inference, the tissue regions are divided in a grid with a fixed stride, and the model perform prediction in a sliding window fashion over the grid. This allows obtaining predictions for the entire tissue regions. During training, we initially performed a balanced random sampling of tiles from the tissue regions for first two epochs; this meant that we alternated between a positive WSI and a negative WSI and selecting an equal number of tiles from each. After the second epoch, we switched into hard mining of tiles, whereby we alternated between a positive WSI and a negative WSI; however, this time performing a sliding window inference on the entire tissue regions and then selecting the top *k* tiles with the highest probabilities for being positive. If the WSI is negative, this effectively selects the tiles most likely to be false positives. The selected tiles were placed in a training subset, and once that subset contained *N* tiles, a training was run whereby the model weights get updated. We used *k* = 8, *N* = 256, and a batch size of 32.

We optimised the model weights by minimising the binary cross-entropy loss using the Adam optimization algorithm Kingma and Ba (2014) with the following parameters: *beta*_1_ = 0.9, *beta*_2_ = 0.999 and a learning rate of 0.001. We applied a learning rate decay of 0.95 every 2 epochs. We used early stopping by tracking the performance of the model on a validation set; this allows stopping the training when no improvement was observed for more than 10 epochs. The model with the lowest validation loss was chosen as the final model.

### 2.4 Software and statistical analysis

The deep learning models were implemented and trained using TensorFlow Abadi et al (2015). AUCs were calculated in python using the scikit-learn package Pedregosa et al (2011) and plotted using matplotlib Hunter (2007). The 95% CIs of the AUCs were estimated using the bootstrap method Efron and Tibshirani (1994) with 1000 iterations.

The true positive rate (TPR) was computed as

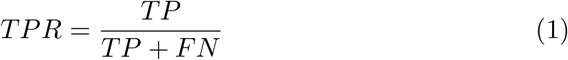

and the false positive rate (FPR) was computed as

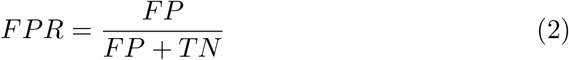

Where TP, FP, and TN represent true positive, false positive, and true negative, respectively. The ROC curve was computed by varying the probability threshold from 0.0 to 1.0 and computing both the TPR and FPR at the given threshold.

### 2.5 Availability of data and material

The datasets generated during and/or analysed during the current study are not publicly available due to specific institutional requirements governing privacy protection but are available from the corresponding author on reasonable request. The datasets that support the findings of this study are available from International University of Health and Welfare, Mita Hospital (Tokyo, Japan) and Kamachi Group Hospitals (Fukuoka, Japan), but restrictions apply to the availability of these data, which were used under a data use agreement which was made according to the Ethical Guidelines for Medical and Health Research Involving Human Subjects as set by the Japanese Ministry of Health, Labour and Welfare, and so are not publicly available. However, the data are available from the authors upon reasonable request for private viewing and with permission from the corresponding medical institutions within the terms of the data use agreement and if compliant with the ethical and legal requirements as stipulated by the Japanese Ministry of Health, Labour and Welfare.

### 2.6 Code availability

To train the classification model in this study we adapted the publicly available TensorFlow training script available at https://github.com/tensorflow/models/tree/master/official/vision/imageclassification.

## 3 Results

### 3.1 Insufficient AUC performance of WSI adenocarcinoma evaluation using existing stomach adenocarcinoma classification model

Prior to the training of multi-organ adenocarcinoma model, we have demonstrated the existing stomach adenocarcinoma classification model Iizuka et al (2020) AUC performance on test sets (Table 3 and 5). Table 6 and Figure 2A show that stomach and colon endoscopic biopsy WSIs and breast TCGA public datasets exhibited high ROC-AUC and low log loss values but not in lung TBLB, breast needle biopsy, and radical lymph node dissection WSIs. Thus, we have trained the models using different WSI number of training sets (Table 2).

**Table 6:** ROC-AUC and log loss results for adenocarcinoma classification on test sets using existing stomach adenocarcinoma classification model

| test sets | Existing stomach adenocarcinoma model |  |
| --- | --- | --- |
|  | ROC-AUC | log loss |
| Stomach endoscopic biopsy | 0.937 [0.918 - 0.953] | 0.450 [0.364 - 0.557] |
| Colon endoscopic biopsy | 0.986 [0.977 - 0.992] | 0.192 [0.142 - 0.252] |
| Lung TBLB | 0.698 [0.665 - 0.726] | 1.807 [1.680 - 1.960] |
| Breast needle biopsy | 0.888 [0.864 - 0.907] | 1.225 [1.111 - 1.329] |
| Lymph node radical dissection | 0.804 [0.771 - 0.832] | 1.940 [1.787 - 2.091] |
| Lung TCGA | 0.797 [0.757 - 0.829] | 1.545 [1.387 - 1.732] |
| Breast TCGA | 0.991 [0.964 - 1.000] | 0.024 [0.012 - 0.039] |

**Fig. 2:**
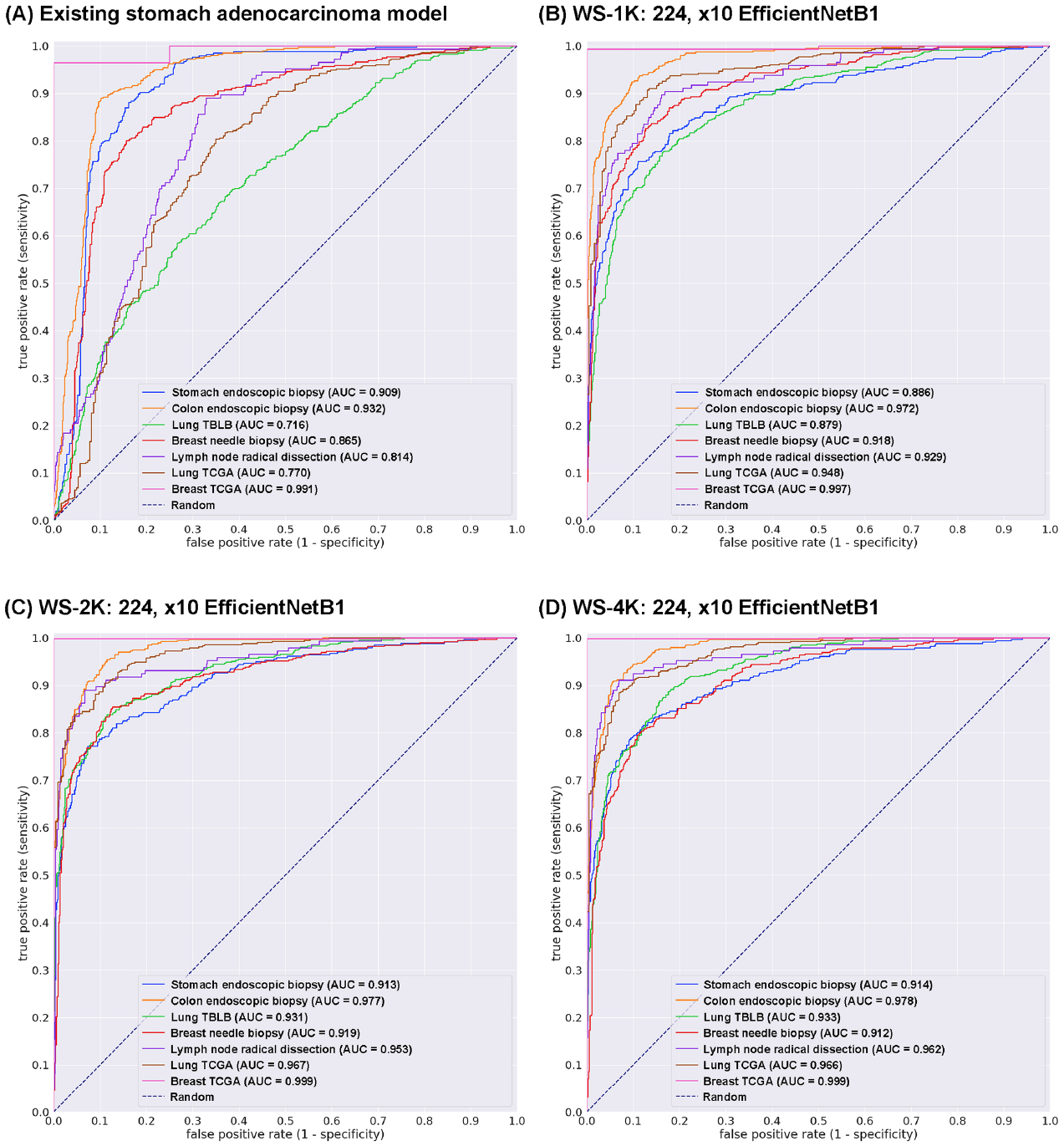
ROC curves with AUCs from four different models (A-D) on the test sets: (A) existing stomach adenocarcinoma classification model and weakly supervised (WS) learning models based on 1K (B), 2K (C), and 4K (D) training sets with tile size 224 px and magnification at x10.

### 3.2 High AUC performance of WSI evaluation of adenocarcinoma histopathology images

We trained models using weakly-supervised (WS) learning which could be used with weak labels (WSI labels) Kanavati et al (2020). We trained using the EfficientNetB1 convolutional neural network (CNN) architecture at magnification x10. The models were applied in a sliding window fashion with input tiles of 224×224 pixels and a stride of 256 (Fig. 1). To train the deep learning models, we used a total of 1,000 (1K), 2,000 (2K), and 4,000 (4K) training set WSIs (Table 2). This resulted in three different models: (1) WS-1K: 224, x10 EfficientNetB1, (2) WS-2K: 224, x10 EfficientNetB1, and (3) WS-4K: 224, x10 EfficientNetB1. We evaluated the models on test sets from domestic hospitals (Table 3) and TCGA public datasets (Table 5). For each test set (stomach endoscopic biopsy, colon endoscopic biopsy, lung TBLB, breast needle biopsy, radical lymph node dissection, lung TCGA, and breast TCGA), we computed the ROC-AUC, log loss, accuracy, sensitivity, and specificity and summarized the results in Table 7 and 8 and Fig. 2B-D. The models trained using 2K and 4K training sets have a higher ROC-AUCs compared to the model trained using 1K and existing stomach adenocarcinoma model (Table 7, Fig. 2). However, there was no obvious difference between the model trained using 2K and 4K training sets (Table 7, Fig. 2C-D). The model (WS-4K: 224, x10 EfficientNetB1) achieved highest ROC-AUC of 0.9993 (CI: 0.9966 - 1.0000) and lowest log loss of 0.0723 (CI: 0.0579 - 0.0903) for adenocarcinoma classification in breast TCGA test sets (Table 7). In test sets from domestic hospitals, the model (WS-4K: 224, x10 EfficientNetB1) achieved very high ROC-AUCs (0.9123 - 0.9776) with low values of log loss (0.203 - 0.437) (Table 7). In all test sets, the model (WS-4K: 224, x10 EfficientNetB1) achieved very high accuracy (0.853 - 0.9987), sensitivity (0.7959 - 0.9986), and specificity (0.8245 - 1.0000) (Table 8). As shown in Fig. 2, Tables 6, 7, and 8, the model (WS-4K: 224, x10 EfficientNetB1) is fully applicable for multi-organ adenocarcinoma classification in wide variety of organs (stomach, colon, lung, breast, and lymph node) WSIs as well as TCGA public WSI dataset. Figures 3, 4, 5, 6, and 7 show representative cases of true positive, true-negative, false positive, and false negative, respectively from using the model (WS-4K: 224, x10 EfficientNetB1).

**Table 7:** ROC-AUC and log loss results for adenocarcinoma classification on test sets using trained models

| WS-1K: 224, x10 EfficientNetB1 |  |  |  |
| --- | --- | --- | --- |
| test sets | ROC-AUC |  | log loss |
| Stomach endoscopic biopsy | 0.886 | [0.864 - 0.912] | 0.415 [0.356 - 0.473] |
| Colon endoscopic biopsy | 0.973 | [0.964 - 0.981] | 0.209 [0.175 - 0.242] |
| Lung TBLB | 0.879 | [0.859 - 0.900] | 0.501 [0.443 - 0.555] |
| Breast needle biopsy | 0.919 | [0.900 - 0.935] | 0.358 [0.315 - 0.404] |
| Lymph node radical dissection | 0.929 | [0.903 - 0.951] | 0.427 [0.380 - 0.486] |
| Lung TCGA | 0.948 | [0.930 - 0.963] | 0.282 [0.238 - 0.333] |
| Breast TCGA | 0.997 | [0.990 - 1.000] | 0.122 [0.097 - 0.150] |

| WS-2K: 224, x10 EfficientNetB1 |  |  |  |
| --- | --- | --- | --- |
| test sets | ROC-AUC |  | log loss |
| Stomach endoscopic biopsy | 0.913 | [0.894 - 0.932] | 0.351 [0.301 - 0.396] |
| Colon endoscopic biopsy | 0.977 | [0.969 - 0.936] | 0.197 [0.167 - 0.226] |
| Lung TBLB | 0.931 | [0.915 - 0.946] | 0.342 [0.300 - 0.386] |
| Breast needle biopsy | 0.919 | [0.901 - 0.936] | 0.371 [0.325 - 0.423] |
| Lymph node radical dissection | 0.953 | [0.939 - 0.978] | 0.228 [0.188 - 0.257] |
| Lung TCGA | 0.967 | [0.955 - 0.977] | 0.246 [0.212 - 0.287] |
| Breast TCGA | 0.999 | [0.996 - 1.000] | 0.146 [0.119 - 0.177] |

| WS-4K: 224, x10 EfficientNetB1 |  |  |  |
| --- | --- | --- | --- |
| test sets | ROC-AUC |  | log loss |
| Stomach endoscopic biopsy | 0.914 | [0.890 - 0.931] | 0.355 [0.315 - 0.404] |
| Colon endoscopic biopsy | 0.978 | [0.970 - 0.984] | 0.203 [0.173 - 0.236] |
| Lung TBLB | 0.933 | [0.917 - 0.946] | 0.437 [0.391 - 0.494] |
| Breast needle biopsy | 0.912 | [0.894 - 0.930] | 0.374 [0.330 - 0.421] |
| Lymph node radical dissection | 0.962 | [0.942 - 0.978] | 0.309 [0.272 - 0.356] |
| Lung TCGA | 0.966 | [0.955 - 0.978] | 0.231 [0.190 - 0.266] |
| Breast TCGA | 0.999 | [0.997 - 1.000] | 0.072 [0.058 - 0.090] |

**Table 8:**
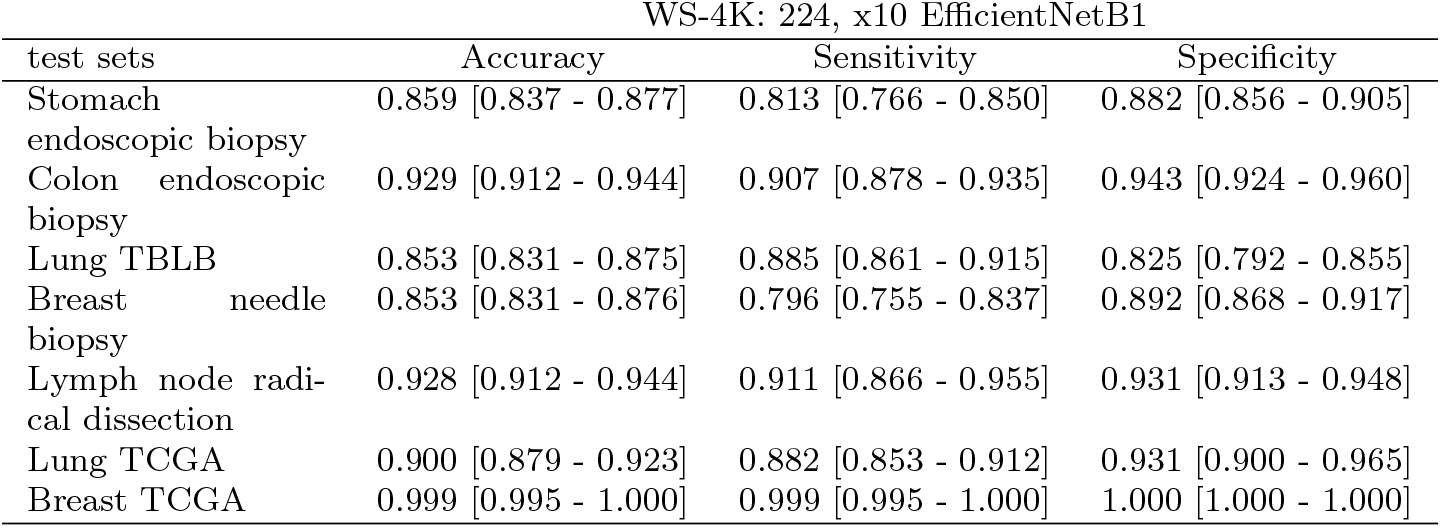
Scores of accuracy, sensitivity, and specificity on test sets using the best model (WS-4K: 224, x10 EfficientNetB1)

**Fig. 3:**
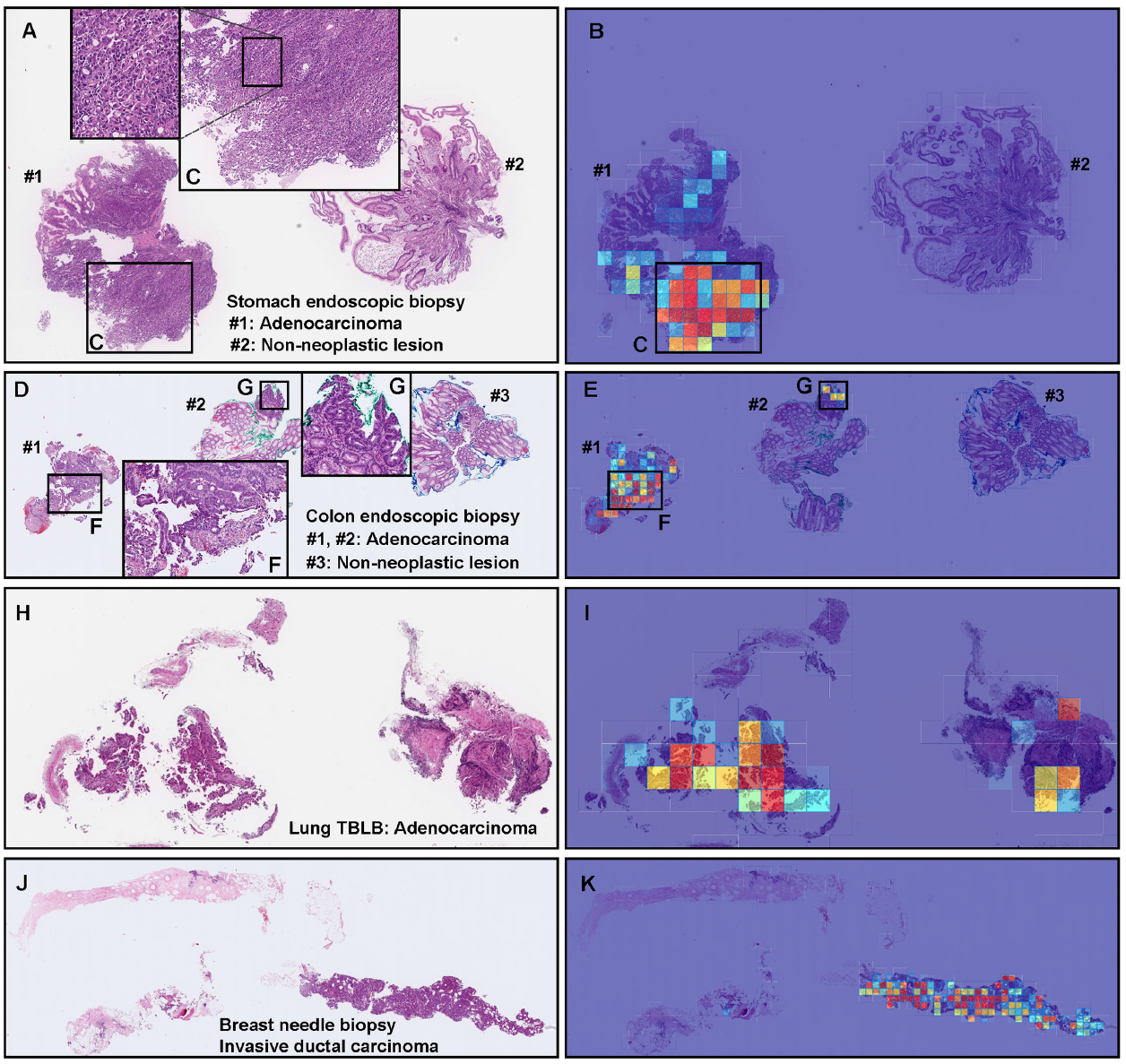
Representative true positive adenocarcinoma classification of stomach, colon, lung, and breast biopsy test cases using the model (WS-4K: 224, x10 EfficientNetB1). In the adenocarcinoma whole slide images (WSIs) of stomach endoscopic biopsy (A), colon endoscopic biopsy (D), lung transbronchial lung biopsy (TBLB) (H), and breast core needle biopsy (J) specimens, the heatmap images show true positive prediction of adenocarcinoma cells (B, E, F, G, I, K) which correspond respectively to H&E histopathology (A, C, D, F, G, H, J). The heatmap images show true negative predictions of non-neoplastic lesion tissue fragments (#2 in (B) and #3 in (E)) and true positive predictions of adenocarcinoma tissue fragments (#1 in (B) and #1-#2 in (E)) which correspond respectively to H&E histopathology of adenocarcinoma area (C, F, G). The heatmap uses the jet color map where blue indicates low probability and red indicates high probability.

**Fig. 4:**
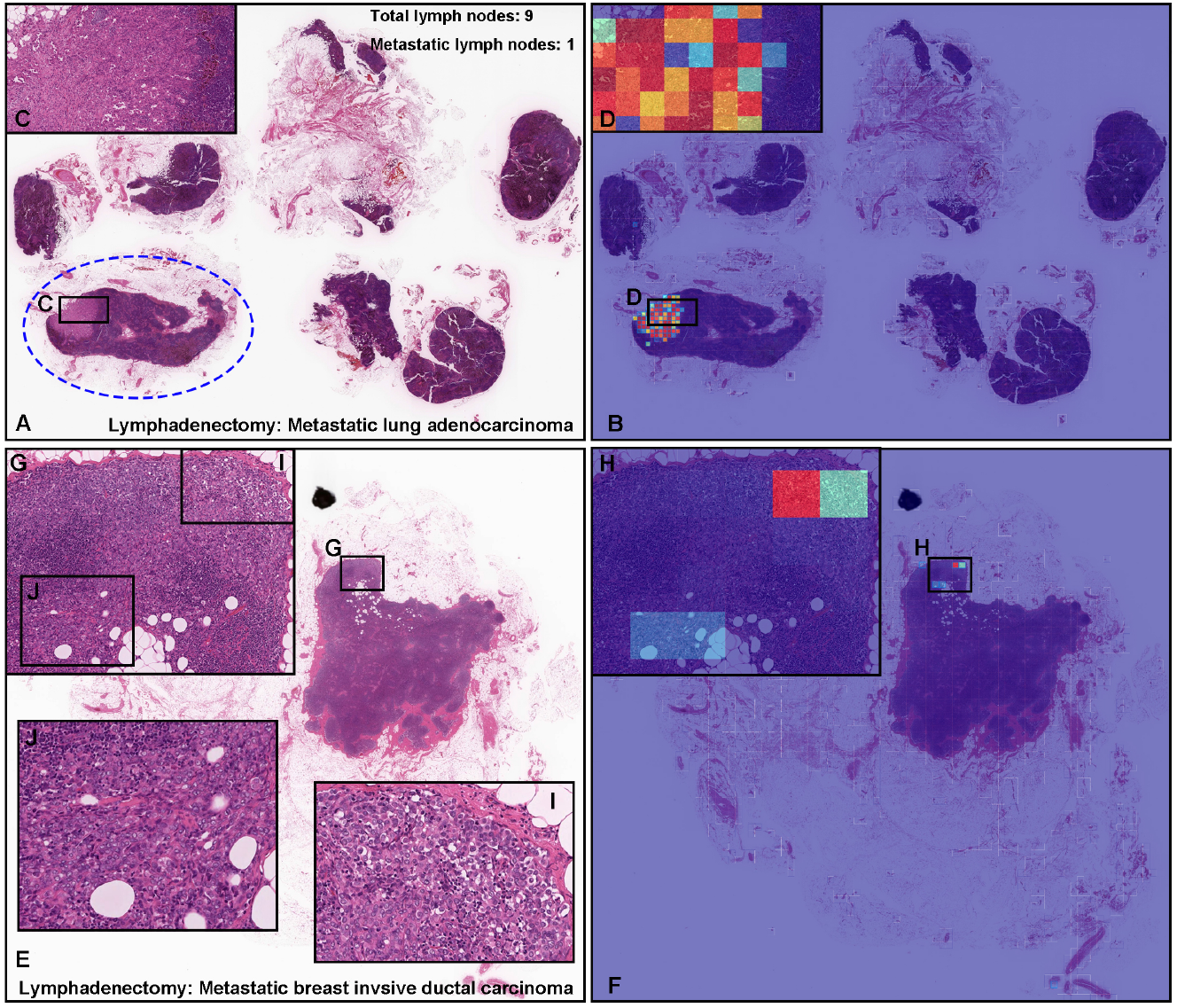
Representative examples of metastatic adenocarcinoma true positive prediction outputs on cases from radical lymph node dissection (lymphadenectomy) test sets using the model (WS-4K: 224, x10 EfficientNetB1). In the metastatic lung adenocarcinoma (A) and breast invasive ductal carcinoma (E) whole slide images (WSIs) of radical lymph node dissection specimens, the heatmap images show true positive prediction of metastatic lung adenocarcinoma (B, D) and breast invasive ductal carcinoma (F, H) cells which correspond respectively to H&E histopathology (A, C, E, G, I, J). According to the histopathological diagnostic report, in (A), only one lymph node (blue dot line circled) was positive for metastatic lung adenocarcinoma (C). The heatmap image (B) shows true positive prediction which was consistent with areas of metastatic lung adenocarcinoma invasion in the same lymph node (D). The heatmap image (B) also shows no positive predictions in the lymph nodes without evidence of cancer metastasis (A). As compared to (A), histopathologically, it was not easy to determine metastatic cancer areas in (E) at low power view. According to the histopathological report, in (E), metastatic breast invasive ductal carcinoma was localized in (G). The heatmap image (F) shows true positive predictions in (H) which are coincided with metastatic carcinoma infiltrating areas (G, I, J). The heatmap uses the jet color map where blue indicates low probability and red indicates high probability.

**Fig. 5:**
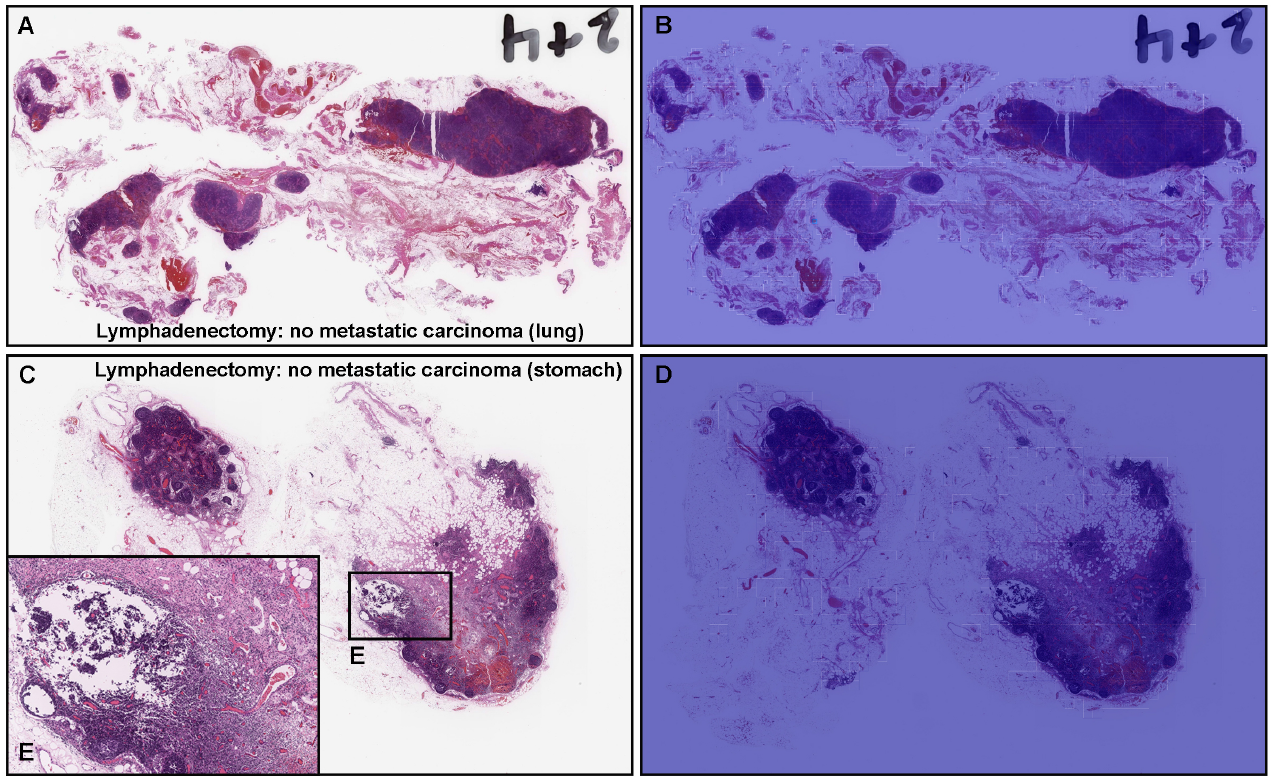
Representative true negative metastatic adenocarcinoma classification of radical lymph node dissection (lymphadenectomy) test sets using the model (WS-4K: 224, x10 EfficientNetB1). Histopathologically, in (A), there were diverse size (small to large) and shape (round to irregular) of lymph nodes without evidence of metastatic adenocarcinoma. The heatmap image (B) shows true negative prediction of metastatic adenocarcinoma. Histopathologically, in (C), there were lymph nodes with lymphadenitis (E) but without evidence of metastatic adenocarcinoma (C, E). The heatmap image (D) shows true negative prediction of metastatic adenocarcinoma. The heatmap uses the jet color map where blue indicates low probability and red indicates high probability.

**Fig. 6:**
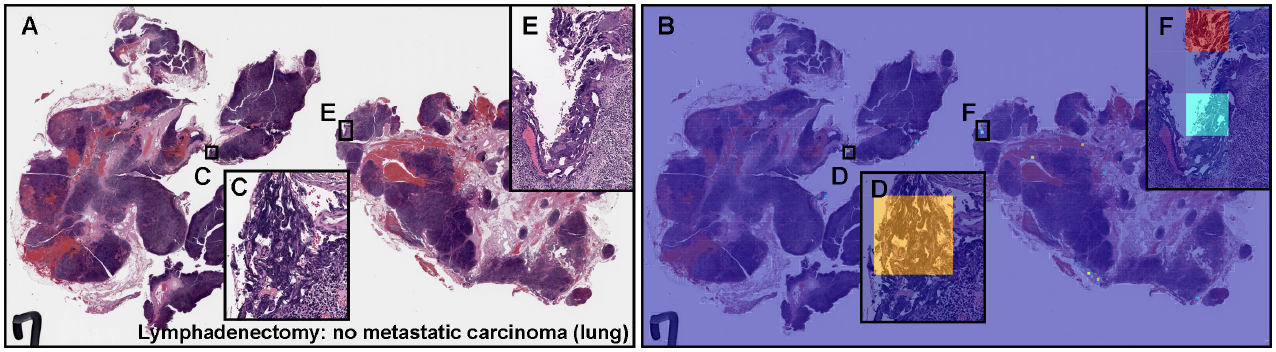
Representative example of metastatic adenocarcinoma false positive prediction outputs on a case from the radical lymph node dissection (lymphadenectomy) test set using the model (WS-4K: 224, x10 EfficientNetB1). Histopathologically, (A) has no sign of metastatic adenocarcinoma. The heatmap image (B) exhibits false positive predictions of adenocarcinoma (D, F) where the tissue consists of dense hematoxylic artifacts induced by crushing during specimen handling procedures (C, E), which most likely is the primary cause of the false positive prediction due to its morphological similarity to adenocarcinoma cells with irregular shaped and dense nuclei. The heatmap uses the jet color map where blue indicates low probability and red indicates high probability.

**Fig. 7:**
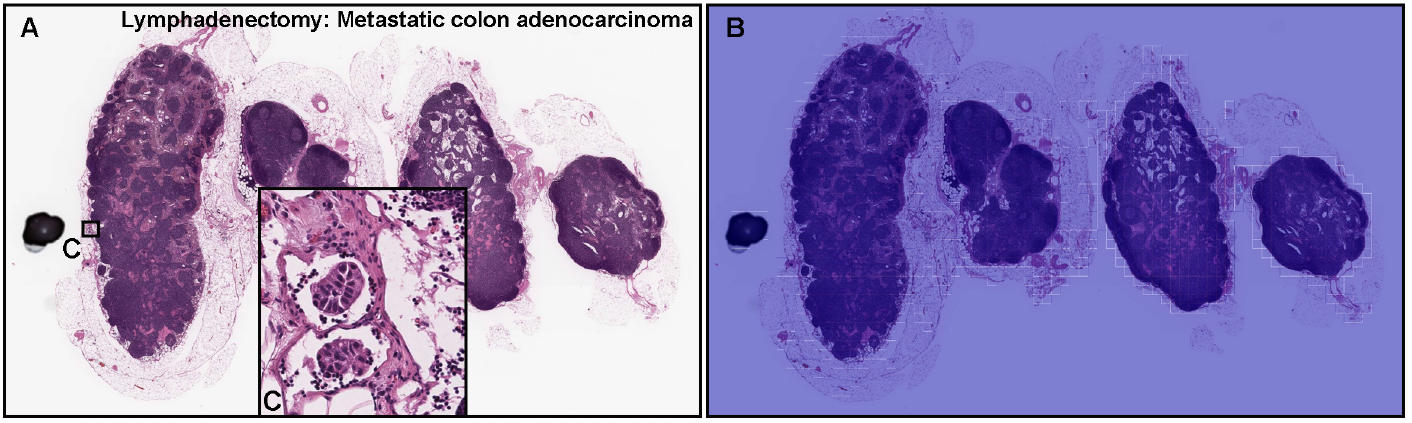
Representative example of metastatic adenocarcinoma false negative prediction output on a case from the radical lymph node dissection (lymphadenectomy) test set using the model (WS-4K: 224, x10 EfficientNetB1). According to the histopathological diagnostic report, this case (A) has metastatic adenocarcinoma foci in (C) but not in other areas. The heatmap image exhibited no positive adenocarcinoma prediction (B). The heatmap uses the jet color map where blue indicates low probability and red indicates high probability.

### 3.3 True positive adenocarcinoma prediction of stomach, colon, lung, and breast biopsy WSIs

Our model (WS-4K: 224, x10 EfficientNetB1) satisfactorily predicted adenocarcinoma in stomach endoscopic biopsy (Fig. 3A, B, C), colon endoscopic biopsy (Fig. 3D, E, F, G), lung TBLB (Fig. 3H, I), and breast needle biopsy (Fig. 3J, K) specimens. Importantly, the heatmap images showed true negative predictions of internal non-neoplastic lesion tissue fragments (#2 in Fig. 3A, B; #3 in Fig. 3D, E; 3H, I, J, K) which were confirmed by surgical pathologists.

### 3.4 True positive adenocarcinoma prediction of radical lymph node dissection (lymphadenectomy) WSIs

A lymphadenectomy (radical lymph node dissection) is a surgical procedure to evaluate evidence of metastatic cancer. In routine histopathological diagnosis, the histopathological inspection of lymph nodes is one of the very important but time-consuming task to avoid the risk of medical oversight. Therefore, in clinical settings, the multi-organ adenocarcinoma model is more useful when performing histopathological diagnosis of lymphadenectomy specimen WSIs. Our model (WS-4K: 224, x10 EfficientNetB1) perfectly predicted metastatic lung adenocarcinoma (Fig. 4A-D) and breast invasive ductal carcinoma (Fig. 4E-J). The heatmap images showed true negative predictions (Fig. 4B) of internal non-neoplastic lymph nodes (Fig. 4A). Importantly, adenocarcinoma localization areas in both metastatic lung adenocarcinoma (Fig. 4C) and breast invasive ductal carcinoma (Fig. 4G) are positively predicted by heatmap images (Fig. 4D, H).

### 3.5 True negative adenocarcinoma prediction of radical lymph node dissection (lymphadenectomy) WSIs

Our model (WS-4K: 224, x10 EfficientNetB1) showed true negative predictions of metastatic adenocarcinoma in lymph nodes without evidence of cancer metastasis (Fig. 5). In Fig. 5A, there were numbers of lymph nodes with broad ranging of size (small to large) and shape (round to irregular) which were not predicted as metastatic lymph nodes (Fig. 5B). Moreover, in Fig. 5C, the lymph node was enlarged due to lymphadenitis (Fig. 5E) but without evidence of metastatic adenocarcinoma which were not predicted as metastatic lymph nodes (Fig. 5D).

### 3.6 False positive adenocarcinoma prediction of radical lymph node dissection (lymphadenectomy) WSIs

Histopathologically, Fig. 6A shows no evidence of metastatic adenocarcinoma. Our model (WS-4K: 224, x10 EfficientNetB1) exhibited false positive predictions of adenocarcinoma (Fig. 6B, D, F). These tissue areas (Fig. 6C, E) showed dense hematoxylic artifacts induced by crushing during specimen handling procedures which could be the primary cause of false positive due to its morphological similarity to irregular shaped and dense nuclei in adenocarcinoma cells.

### 3.7 False negative adenocarcinoma prediction of radical lymph node dissection (lymphadenectomy) WSIs

In Fig. 7A, histopathologically, only two metastatic colon adenocarcinoma foci were observed in the left-most lymph node (Fig. 7C). After double checking two independent pathologists, there were no more metastatic adenocarcinoma cells in Fig. 7A. However, the heatmap image did not predict any adenocarcinoma cells (Fig. 7B).

## 4 Discussion

In the present study, we trained multi-organ deep learning models for the classification of adenocarcinoma in WSIs using weakly-supervised learning. The models were trained on WSIs obtained from four medical institutions and were then applied on multi-organ test sets obtained from five medical institutions and publicly available TCGA datasets to demonstrate the generalization of the model on unseen data. The deep learning model (WS-4K: 224, x10 EfficientNetB1) achieved ROC-AUCs in the range of 0.91-0.99.

So far, we have been investigating adenocarcinoma classification on histopathological WSIs in diverse organs (e.g., stomach Iizuka et al (2020); Kanavati et al (2021a); Kanavati and Tsuneki (2021b), colon Iizuka et al (2020); Tsuneki and Kanavati (2021), lung Kanavati et al (2020, 2021b), and breast Kanavati and Tsuneki (2021a); Kanavati et al (2022)). These models are specific to each organ, and versatile adenocarcinoma histopathological classification model(s) which can be applied in multi-organ have not been developed to date. The global adenocarcinoma classification model in multi-organ may play key roles in first-screening processes especially radical lymph node dissection specimens which consist of a large number of lymph nodes in a single WSI in routine pathological diagnosis in the clinical laboratories.

Prior to the training, we have demonstrated the versatility of the existing models. For example, the existing stomach adenocarcinoma classification model Iizuka et al (2020) exhibited scores of high ROC-AUC and low log loss for the stomach and colon endoscopic biopsy test sets, but not for the lung, breast, and lymph node test sets (Table 6). Therefore, we have trained the deep learning models from scratch by the weakly-supervised learning approach in this study.

We have collected histopathological H&E stained specimens from as many medical institutions as possible to ensure diversities of histopathological variability and specimen quality in training sets (Table 1). In the training sets, we did not include radical lymph node dissection specimens because we would like to train the model based on the primary organs and predict metastatic adenocarcinoma in lymph nodes. In all training sets (1K, 2K, and 4K), WSIs from each organ (stomach, colon, lung, and breast) were equally distributed (Table 2).

In this study, we showed that it was possible to exploit the use of a moderate size training sets of 2,000 (2K) and 4,000 (4K) WSIs to train deep learning models using a weakly-supervised learning, and we have obtained high ROCAUC performance on primary organ (stomach, colon, lung, and breast) and radical lymph node dissection test sets as well as public TCGA datasets (lung and breast), which is highly promising in terms of the generalisation performance of our models to classify adenocarcinoma in multi-organs. Using the weakly-supervised learning method allowed us to train on our datasets and obtain high performance without manually performed annotations. This means that it is possible to train a very high performance model for any type of cancer classification in multi-organ without having to have detailed cellular level or rough annotations or requiring an extremely large number of WSI. We have demonstrated the usefulness of weakly-supervised learning approach for lung carcinoma classification Kanavati et al (2020). Importantly, there were no significant difference in ROC-AUC and log loss results between 2K and 4K training sets, meaning that small number (total 2,000 WSIs) of training datasets were enough for adenocarcinoma classification in multi-organ.

Our model satisfactorily predicted adenocarcinoma areas not only in primary organs (stomach, colon, lung, and breast) (Fig. 3) but also in radical lymph node dissection specimens (Fig. 4). In routine histopathological diagnosis, inspecting cancer metastasis in lymph nodes is laborious because usually there are a lot of lymph nodes with wide variety of sizes and shapes in glass slides. Our model can localise the prediction of adenocarcinoma invasion and visualise them as heatmap images (Fig. 4) which would be a great tool for primary screening or double-check purpose in clinical workflow in laboratories. Importantly, our model can evaluate adenocarcinoma-free (non-metastatic) lymph nodes (Fig. 5) which reflected high specificity (0.931) (Table 8). This is an important finding to apply our model in clinical workflow. As for the false positive and false negative predictions, certain trends were observed. The dense hematoxylic artifacts induced by crushing during specimen handling procedures were observed as the primary causes of the false positive prediction which have morphological similarities to adenocarcinoma cell clusters with irregular shaped and dense nuclei (Fig. 6). In the next step for further training, it would be best to collect appropriate number of false positive and false negative cases and perform active learning.

## 5 Acknowledgements

We are grateful for the support provided by Dr. Shin Ichihara at Department of Surgical Pathology, Sapporo Kosei General Hospital (Sapporo, Japan); Dr. Makoto Abe at Department of Pathology, Tochigi Cancer Center (Tochigi, Japan); Dr. Shigeo Nakano at Kamachi Group Hospitals (Fukuoka, Japan); Professor Takayuki Shiomi at Department of Pathology, Faculty of Medicine, International University of Health and Welfare (Tokyo, Japan); Dr. Ryosuke Matsuoka at Diagnostic Pathology Center, International University of Health and Welfare, Mita Hospital (Tokyo, Japan). We thank pathologists who have been engaged in reviewing cases and clinicopathological discussion for this study.

## 6 Compliance with Ethical Standards

The experimental protocol was approved by the ethical board of International University of Health and Welfare (No. 19-Im-007) and Kamachi Group Hospitals (No. 173). All research activities complied with all relevant ethical regulations and were performed in accordance with relevant guidelines and regulations in the all hospitals mentioned above. Informed consent to use histopathological samples and pathological diagnostic reports for research purposes had previously been obtained from all patients prior to the surgical procedures at all hospitals, and the opportunity for refusal to participate in research had been guaranteed by an opt-out manner.

## 9 Contributions

M.T. and F.K. contributed equally to this study; F.K. and M.T. designed the studies, performed experiments and analyzed the data; F.K. and M.T. wrote the manuscript; M.T. supervised the project. All authors reviewed and approved the final manuscript.

## References

Abadi M, Agarwal A, Barham P, et al (2015) TensorFlow: Large-scale machine learning on heterogeneous systems. URL https://www.tensorflow.org/, software available from tensorflow.org

Bejnordi BE, Veta M, Van Diest PJ, et al (2017) Diagnostic assessment of deep learning algorithms for detection of lymph node metastases in women with breast cancer. Jama 318(22):2199–2210

Campanella G, Hanna MG, Geneslaw L, et al (2019) Clinical-grade computational pathology using weakly supervised deep learning on whole slide images. Nature medicine 25(8):1301–1309

Coudray N, Ocampo PS, Sakellaropoulos T, et al (2018) Classification and mutation prediction from non–small cell lung cancer histopathology images using deep learning. Nature medicine 24(10):1559–1567

Efron B, Tibshirani RJ (1994) An introduction to the bootstrap. CRC press

Gertych A, Swiderska-Chadaj Z, Ma Z, et al (2019) Convolutional neural networks can accurately distinguish four histologic growth patterns of lung adenocarcinoma in digital slides. Scientific reports 9(1):1483

Goode A, Gilbert B, Harkes J, et al (2013) Openslide: A vendor-neutral software foundation for digital pathology. Journal of pathology informatics 4

Hou L, Samaras D, Kurc TM, et al (2016) Patch-based convolutional neural network for whole slide tissue image classification. In: Proceedings of the IEEE Conference on Computer Vision and Pattern Recognition, pp 2424– 2433

Hunter JD (2007) Matplotlib: A 2d graphics environment. Computing in Science & Engineering 9(3):90–95. https://doi.org/10.1109/MCSE.2007.55

Iizuka O, Kanavati F, Kato K, et al (2020) Deep learning models for histopathological classification of gastric and colonic epithelial tumours. Scientific reports 10(1):1–11

Kanavati F, Tsuneki M (2021a) Breast invasive ductal carcinoma classification on whole slide images with weakly-supervised and transfer learning. bioRxiv

Kanavati F, Tsuneki M (2021b) A deep learning model for gastric diffuse-type adenocarcinoma classification in whole slide images. arXiv preprint arXiv:210412478

Kanavati F, Tsuneki M (2021c) Partial transfusion: on the expressive influence of trainable batch norm parameters for transfer learning. arXiv preprint arXiv:210205543

Kanavati F, Toyokawa G, Momosaki S, et al (2020) Weakly-supervised learning for lung carcinoma classification using deep learning. Scientific reports 10(1):1–11

Kanavati F, Ichihara S, Rambeau M, et al (2021a) Deep learning models for gastric signet ring cell carcinoma classification in whole slide images. Technology in Cancer Research & Treatment 20:15330338211027,901

Kanavati F, Toyokawa G, Momosaki S, et al (2021b) A deep learning model for the classification of indeterminate lung carcinoma in biopsy whole slide images. Scientific Reports 11(1):1–14

Kanavati F, Ichihara S, Tsuneki M (2022) A deep learning model for breast ductal carcinoma in situ classification in whole slide images. Virchows Archiv pp 1–14

Kingma DP, Ba J (2014) Adam: A method for stochastic optimization. arXiv preprint arXiv:14126980

Korbar B, Olofson AM, Miraflor AP, et al (2017) Deep learning for classification of colorectal polyps on whole-slide images. Journal of pathology informatics 8

Kraus OZ, Ba JL, Frey BJ (2016) Classifying and segmenting microscopy images with deep multiple instance learning. Bioinformatics 32(12):i52–i59

Litjens G, Sánchez CI, Timofeeva N, et al (2016) Deep learning as a tool for increased accuracy and efficiency of histopathological diagnosis. Scientific reports 6:26,286

Luo X, Zang X, Yang L, et al (2017) Comprehensive computational pathological image analysis predicts lung cancer prognosis. Journal of Thoracic Oncology 12(3):501–509

Madabhushi A, Lee G (2016) Image analysis and machine learning in digital pathology: Challenges and opportunities. Medical Image Analysis 33:170– 175

Otsu N (1979) A threshold selection method from gray-level histograms. IEEE transactions on systems, man, and cybernetics 9(1):62–66

Pedregosa F, Varoquaux G, Gramfort A, et al (2011) Scikit-learn: Machine learning in Python. Journal of Machine Learning Research 12:2825–2830

Saltz J, Gupta R, Hou L, et al (2018) Spatial organization and molecular correlation of tumor-infiltrating lymphocytes using deep learning on pathology images. Cell reports 23(1):181–193

Sung H, Ferlay J, Siegel RL, et al (2021) Global cancer statistics 2020: Globocan estimates of incidence and mortality worldwide for 36 cancers in 185 countries. CA: a cancer journal for clinicians 71(3):209–249

Tan M, Le Q (2019) Efficientnet: Rethinking model scaling for convolutional neural networks. In: International Conference on Machine Learning, PMLR, pp 6105–6114

Tsuneki M, Kanavati F (2021) Deep learning models for poorly differentiated colorectal adenocarcinoma classification in whole slide images using transfer learning. Diagnostics 11(11):2074

Wei JW, Tafe LJ, Linnik YA, et al (2019) Pathologist-level classification of histologic patterns on resected lung adenocarcinoma slides with deep neural networks. Scientific reports 9(1):1–8

Yu KH, Zhang C, Berry GJ, et al (2016) Predicting non-small cell lung cancer prognosis by fully automated microscopic pathology image features. Nature communications 7:12,474

